# Asymptomatic carotid artery stenosis, cognitive decline and dementia: a prospective population-based cohort study

**DOI:** 10.64898/2026.08.18.26360680

**Authors:** Camiel V.J. Box, Amos C. Pomp, Qingling Yu, Maryam Kavousi, M. Kamran Ikram, Aad van der Lugt, Daniel Bos, Frank J. Wolters

## Abstract

**Background:** Asymptomatic carotid artery stenosis (ACAS) increases the risk of stroke, and is associated with cognitive decline. This association might be driven by underlying atherosclerotic disease instead of the stenosis itself, which could explain why studies on the cognitive benefits of carotid revascularisation are inconclusive.

**Methods:** Between 2007-2012, dementia-free participants of the population-based Rotterdam Study underwent carotid ultrasound, and additional carotid MRI if intimal media thickness was >2.5mm. All participants underwent repeated cognitive assessments and were followed for dementia until January 2022. We determined the effect of ACAS and plaque without stenosis on all-cause dementia using multivariable Cox models, and on change in cognition (g-factor) using multivariable linear mixed-effects models.

**Results:** Of 4267 participants (mean age 67.5 years, 55.5% women), 483 (11.3%) had plaque without stenosis, 989 (23.2%) had 1-49% stenosis, 107 (2.5%) had 50-99% stenosis, and 15 (0.4%) had occlusion. During a mean follow-up of 9.7 years, 391 participants developed dementia. Compared to individuals without carotid atherosclerosis, risk of dementia was increased in the presence of plaque without stenosis (HR: 1.39 [95%CI: 1.05−1.83]) and occlusion (HR: 4.61 [1.82−11.66]), but not with stenosis (HR 1-49% stenosis: 1.08 [0.84−1.39]; 50-99% stenosis: 1.18 [0.70−1.99]). Neither carotid plaques nor stenosis affected cognitive decline. Results did not differ consistently by plaque characteristics.

**Conclusion:** Risk of dementia was increased with asymptomatic carotid artery plaque and occlusion, but not significantly with 50-99% stenosis. These results are in line with detrimental effects of generalised atherosclerotic disease and severe haemodynamic impairment on cognitive decline and dementia risk.

**KEY MESSAGES:** *What is already known on this topic:* We conducted a systematic review and meta-analysis on the effects of carotid atherosclerosis and revascularisation on dementia and cognition, which is published separately. We found consistent evidence for an association between multiple measures of carotid atherosclerosis and dementia. However, no studies investigated the distinct effects of atherosclerosis, stenosis, and occlusion. Interventional studies did not provide robust evidence for a positive effect of carotid revascularisation on cognition or dementia.

*What this study adds:* In this large prospective cohort study, we found that risk of dementia was increased with carotid plaque without stenosis and full occlusion, but no significant effect was found with mild, moderate or severe stenosis. Effects of carotid occlusion on dementia may manifest only with severe reduction in blood flow not generally observed with stenosis alone.

*How this study might affect research, practice or policy:* Reducing the burden of atherosclerosis – and optimal management of its risk factors–warrants attention in individuals with ACAS for prevention of dementia. Our findings further caution about haemodynamic consequences on cognition in patients with complete occlusion, but do not support revascularisation for the benefit of cognition in most cases with ACAS.

## INTRODUCTION

Carotid artery atherosclerosis is an important cause of ischemic stroke, with higher degrees of stenosis reflecting higher stroke risk.^1^ Urgent carotid endarterectomy or stenting after TIA or stroke is a cornerstone of secondary stroke prevention in patients with moderate to severe carotid stenosis.^2^ Yet, whether revascularisation should also be offered to patients without a history of TIA or stroke is heavily debated,^3^ ^4^ with mixed evidence from the recent ESCT-2 and CREST-2 trials in terms of stroke prevention.^5–8^ Asymptomatic carotid artery stenosis (ACAS) with ≥50% luminal narrowing is present in 0.6% of the general population at age 50-59, increasing to 6.2% after age 80, rendering clear directions about its management relevant to a potentially sizeable group of older adults.^9^

Benefits of revascularisation could extend beyond stroke prevention if ACAS also contributes to cognitive decline and dementia. Such effects on cognition beyond the direct effects of stroke are plausible, in view of contributions of reduced cerebral perfusion and subclinical (micro)infarction to dementia risk.^10–12^ Yet, direct evidence for a causal contribution of ACAS to dementia risk is scarce.^13^ In a concurrent systematic review and meta-analysis, we show that multiple measures of carotid atherosclerosis, including stenosis, were associated with dementia risk. However, excluding the present study, only two other longitudinal observational cohort studies were included in the meta-analysis on the effect of ≥50% stenosis on dementia.^14^ ^15^ Furthermore, two important caveats arose from our evidence synthesis on the effect of stenosis on dementia: (1) studies used stenosis thresholds such as 50% to improve clinical relevance but by doing so included full occlusion, even though these cannot be intervened on; (2) no study examined stenosis and the underlying atherosclerotic burden within a single model, leaving unresolved whether the observed association between stenosis and dementia risk is driven by the stenosis itself, which could be resolved by intervention, or by the broader atherosclerotic disease process.

Moreover, stroke risk in individuals with ACAS varies substantially by plaque characteristics, notably intraplaque haemorrhage,^2^ ^16–18^ but whether plaque characteristics affect cognitive decline and dementia risk is undetermined.

We therefore investigated the effects of asymptomatic carotid artery stenosis, as compared to non-stenotic atherosclerosis, on cognitive decline and the long-term risk of dementia in a population-based study. We further explored whether these risks differ by plaque characteristics.

## METHODS

The Strengthening the Reporting of Observational Studies in Epidemiology (STROBE) guideline was used to guide reporting (Table S1).

### Study design and participants

This study was embedded in the Rotterdam Study, a prospective population-based cohort study of 17,931 residents of the Ommoord district in Rotterdam, the Netherlands.^19^ Details of the Rotterdam Study have been described previously.^19^ In brief, the original cohort was recruited in 1990, including all persons aged 55 years and older, and has been expanded thrice since, in 2000, 2006 and 2016, to include all residents who reached age 40 or moved into the study area. The overall response figure was 65%. All participants are examined every 3 to 5 years at a dedicated research centre, including extensive cognitive screening and routine brain MRI.

For the current analyses, we considered 8795 consecutive dementia-free participants who underwent carotid artery ultrasonography between 2002 and 2014. Of 4835 (55%) participants with a carotid intima-media thickness (IMT) >2.5mm on ultrasound, a random subsample was invited for carotid MRI to further characterise stenosis and plaque characteristics. Of 2446 invitees, 1680 (69%) successfully completed carotid MRI between 2007 and 2012, whereas 161 (7%) had a contraindication for MRI, and 605 (25%) declined or did not undergo MRI for other reasons. To ensure comparability of groups with and without carotid artery atherosclerosis (i.e. avoid healthy volunteer bias with respect to MRI participation), we included only the 2674 of 3960 (68%) reference participants with IMT ≤2.5mm who underwent brain MRI as part of the Rotterdam Study in the same time period. Finally, we excluded 9 participants with a history of carotid endarterectomy prior to study baseline, and 79 participants with a history of ipsilateral ischemic stroke or TIA, leaving a total 4267 participants for analysis (Figure 1).

**Figure 1:**
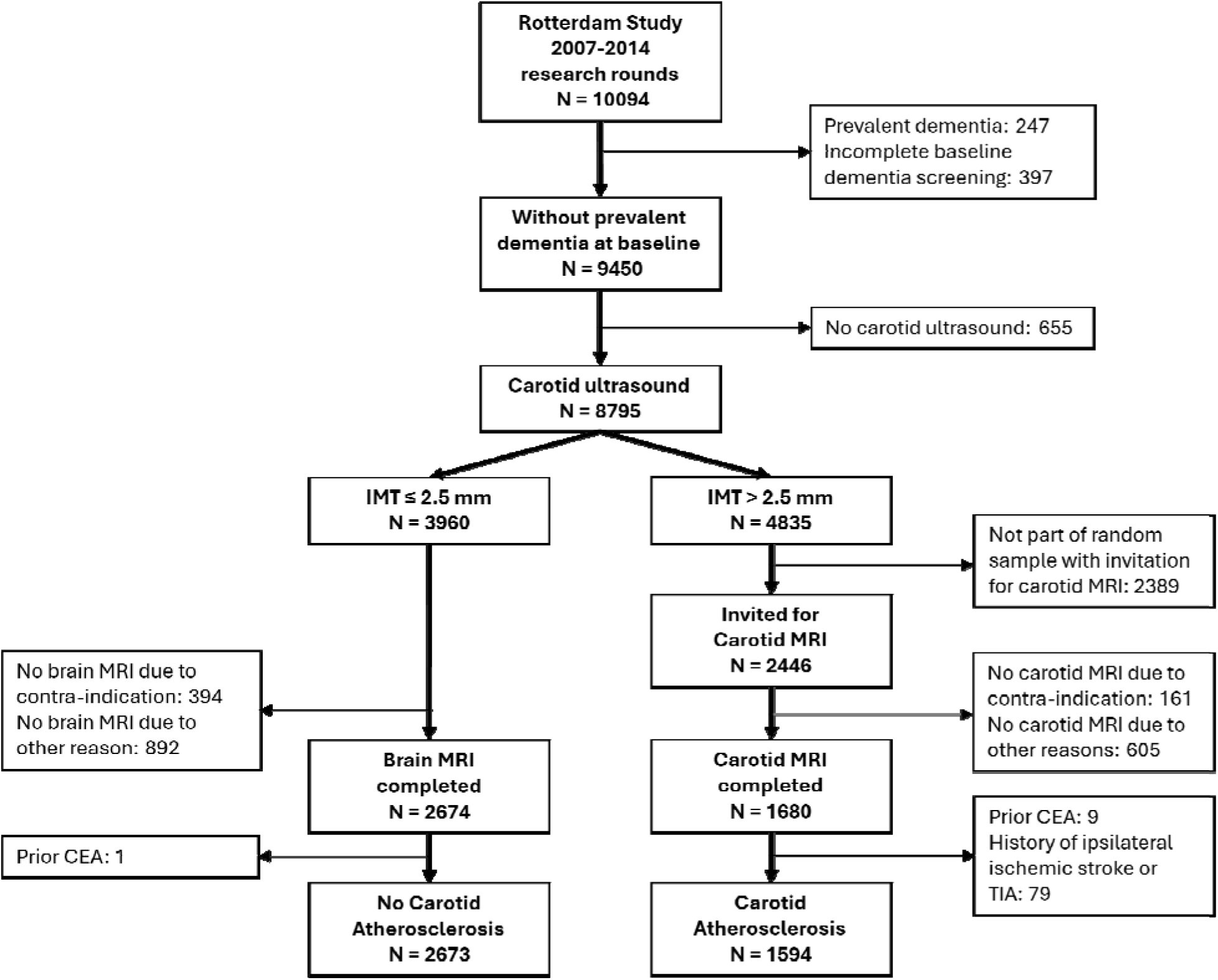
Inclusion flowchart. Legend: CEA = carotid endarterectomy; IMT = intimal medial thickness

The Rotterdam Study has been approved by the Medical Ethics Committee of the Erasmus MC (registration number MEC 02.1015) and by the Dutch Ministry of Health, Welfare and Sport (Population Screening Act WBO, license number 1071272-159521-PG). The Rotterdam Study Personal Registration Data collection is filed with the Erasmus MC Data Protection Officer under registration number EMC1712001. The Rotterdam Study has been entered into the Dutch Trial Register (NTR; https://onderzoekmetmensen.nl) and into the WHO International Clinical Trials Registry Platform (https://www.who.int/clinical-trials-registry-platform, search portal https://trialsearch.who.int/) under shared catalogue number NL6645/NTR6831. All participants provided written informed consent.

### Carotid artery atherosclerosis

A detailed protocol of carotid ultrasonography and carotid MRI in the Rotterdam Study has been published elsewhere.^20^ ^21^ In short, IMT was measured on ultrasonography as the maximum distance between the near and far walls of the common carotid artery, carotid bifurcation, and internal carotid artery on both sides. MRI of the carotid arteries was performed on a 1.5 T scanner (GE Healthcare, Milwaukee, Wisconsin) and rated by trained research physicians, supervised by an experienced neuroradiologist. Rating was blinded to all participant characteristics. Degree of luminal stenosis was quantified according to the NASCET criteria on the proton density weighted fast spin echo sequence,^22^ and plaques were visually rated for the presence of intraplaque haemorrhage, lipid core, and calcification. Intra- and interobserver variability were good, with Cohen’s κ values ranging from 0.77 to 0.88 for stenosis degree, and 0.86 to 0.95 for plaque characteristics.^21^ For the main analyses, participants were divided according to presence of carotid plaque and degree of stenosis: no carotid atherosclerosis, plaque without stenosis, 1-49% stenosis, 50-99% stenosis, and 100% stenosis (i.e. occlusion).^21^ Only in case of ≥50% stenosis, participants and their general practitioners were informed by a study physician.

### Cognitive assessment, and ascertainment of dementia and death

The primary outcome of the study was all-cause dementia, which was assessed through periodic in-person assessments at each research centre visit and continuous linkage with medical records. Those with a Mini Mental State Examination score below 26 or Geriatric Mental Schedule organic level above 0 were invited for anamnesis, informant interview, and further cognitive assessment with the Cambridge Examination for Mental Disorders in the Elderly (CAMDEX). Moreover, all participants underwent a routine cognitive assessment, as detailed below. Along with information from medical records, all results were reviewed by a consensus panel led by a consultant neurologist to determine final diagnoses based on standard diagnostic criteria, including the DSM-III-R for dementia. Dementia ascertainment was blinded to carotid imaging results. Mortality was ascertained through linkage with municipal administration. Follow-up was done until 1^st^ January 2022 for dementia or death, and participants were censored at time of dementia, death, carotid endarterectomy, carotid stenting, or loss to follow-up, whichever came first. Follow-up for dementia was complete for 41,251 of 42,081 potential person-years (98%).

For the secondary outcome of cognition, we derived a global cognition measure (g-factor) from a cognitive test battery administered during each research centre visit, including the Stroop,^23^ verbal fluency (animal naming),^24^ the letter-digit substitution,^25^ the word-learning,^26^ and the Purdue pegboard test. We calculated the g-factor using principal component analysis, with the first component explaining roughly 50% of variance in cognitive performance. Higher standardised values of the g-factor indicate better cognitive performance. To explore domain-specific effects, we also assessed each of the cognitive tests separately. We excluded 78 participants who only had cognitive assessment more than 3 years before imaging. The median absolute time between the first cognitive assessment and carotid artery imaging (ultrasound or MRI) was 1.4 months (IQR: 0.6; 3.7). Overall, we included 4178 (98%) participants who participated in detailed cognitive assessment, for a total of 8228 rounds of cognitive testing, of which 1650 (20.1%) observations were missing in part (i.e. participants completed the MMSE and at least one, but not all, of the five other tests). For computation of the g-factor, we imputed missing scores using random forest imputation based on age, educational attainment, MMSE score, and scores of completed cognitive test(s) if participants had at least one available cognitive test result and a MMSE score.

### Other clinical characteristics

History and occurrence of stroke and TIA were determined through a combination of participant interview and medical records review, and all potential cases were discussed in a consensus panel led by a vascular neurologist. If the side of stroke or TIA was uncertain, we considered it ipsilateral to ensure a population with asymptomatic stenosis.

Information regarding age, sex, highest attained education (four categories according to UNESCO classification),^27^ smoking habits (never, former, current), and medication use (antihypertensives, antithrombotic and lipid lowering) was reported by participants during interviews. Weight and height were measured to calculate body mass index (BMI) in kg/m^2^. Systolic and diastolic blood pressure were measured twice in sitting position on the right arm using a random-zero sphygmomanometer; the mean of two readings was used for analysis. Fasting blood samples were taken to measure glucose, creatine, total cholesterol, and high-density lipoprotein (HDL) cholesterol. Estimated glomerular filtration rates (eGFR) were calculated from creatinine using the CKD-EPI formula. Diabetes was defined as fasting glucose >6.9mmol/L, or the use of antidiabetic medication. *APOE*-ε4 allele carriership was determined using PCR on coded DNA samples, or biallelic TaqMan assays (TaqMan Gene Expression Assays; Thermo Fisher Scientific, Waltham, MA) (rs7412 and rs429358); if PCR or Taqman were unavailable (<7%), we used Haplotype Reference Consortium imputations. All investigations took place at time of carotid ultrasound. As carotid MRI was performed during an additional, dedicated research visit, the interview and measurements of other variables occurred a median of 7.0 months (interquartile range [IQR]: 3.0–44.3) before the time of carotid MRI.

### Statistical analyses

Missing covariate data (education, smoking, BMI, blood pressure, diabetes, and medication use all <1%, cholesterol 1.9%, *APOE*-genotype 4.3%, and eGFR 8.1%) were imputed using five-fold multivariate imputation by chained equation. We calculated age-specific prevalence of 50-99%, 70-99%, and 100% stenosis but corrected for probability of being invited for the carotid MRI for those with IMT >2.5mm (50.4%).

Relative risks of dementia in different exposure groups were quantified as hazard ratios (HR) with 95% confidence intervals (95%CI) from three cause-specific Cox models: (1) crude, (2) adjusted for age, age^2^ and sex, and (3) adjusted additionally for education, smoking status, BMI, systolic and diastolic blood pressure, total and HDL cholesterol, eGFR, diabetes prevalence, *APOE*-e4 carriership, use of antithrombotic, antihypertensive, and lipid lowering medication, and a history of contralateral ischemic stroke. Using similar models, we determined relative risks with carotid plaque characteristic among people with carotid atherosclerosis. We repeated analyses for the outcome of mortality. Schoenfeld residuals and plots of scaled Schoenfeld residuals over time showed no violation of the proportional hazards assumption (Figure S1).

Next, we determined the difference in baseline cognition and cognitive trajectories over time between carotid atherosclerosis groups using linear mixed models with random intercepts, random slopes, restricted maximum likelihood, and an unstructured covariance matrix. Models were adjusted similarly as mentioned earlier, plus a quadratic term for age and an interaction between age and follow-up time. Residual and Q-Q plots are displayed in Figure S2. All scores were standardised and Stroop scores were inverted to ensure higher scores represent a better outcome.

Finally, we performed sensitivity analyses, (1) stratified by sex, (2) by age (<75 and ≥75 years), (3) with separate coefficients for left-sided and right-sided stenosis, (4) censoring at time of ischemic stroke to investigate a possible mediating effect of stroke, (5) with dichotomous exposure variables for atherosclerosis vs no atherosclerosis, ≥50% stenosis vs <50% stenosis, and ≥70% stenosis vs <70% stenosis, and (6) with the same dichotomous exposure variables but excluding occlusion.

Analyses were performed using R Statistical Software (v4.3.3; packages “mice”, “missForest”, “nlme”, and “survival”). Alpha was set at 0.05.

## RESULTS

Mean age of participants was 67.5 (SD: 9.9) years, and 55.5% were female (Table 1). Of all 4267 included participants, 2673 (62.6%) had no signs of carotid artery atherosclerosis, 483 (11.3%) had a plaque without stenosis, 989 (23.2%) a stenosis of 1-49%, 107 (2.5%) a stenosis of 50-99%, and 15 (0.4%) had an occlusion. Prevalence of 50-99% stenosis increased with age, from 0.5% (95%CI: 0–1.7%) at age 45-49, to 5.0% (2.6–7.4) in those aged 80 years or older (Figure S3). For stenosis 70-99%, prevalence ranged from estimated 0% at age 45-49 to 1.3% (0.04–2.5%) after age 80. (Figure S3). Participants with carotid atherosclerosis or stenosis were older, more often male and generally had a worse cardiovascular risk profile (Table 1).

**Table 1.** Baseline characteristics of the study population stratified by carotid disease category.

|  | Total<br>population (N<br>= 4267) | Without carotid<br>atherosclerosis<br>(N = 2673) | Plaque, no<br>stenosis<br>(N = 483) | 1-49%<br>stenosis<br>(N = 989) | 50-99%<br>stenosis<br>(N = 107) | Occlusion<br>(N = 15) |
| --- | --- | --- | --- | --- | --- | --- |
| Age, mean (SD) | 67.5 (9.9) | 64.6 (9.1) | 72.7 (8.9) | 72.1 (9.2) | 74.5 (9.3) | 73.8 (6.5) |
| Female sex, n (%) | 2370 (55.5) | 1640 (61.4) | 227 (47.0) | 453 (45.8) | 47 (43.9) | 3 (20.0) |
| Education, n (%) |  |  |  |  |  |  |
| Primary only | 382 (9.0) | 209 (7.8) | 39 (8.1) | 119 (12.0) | 14 (13.1) | 1 (6.7) |
| Lower/intermediate general, lower vocational | 1595 (37.4) | 986 (36.9) | 197 (40.8) | 367 (37.1) | 40 (37.4) | 5 (33.3) |
| Intermediate vocational, higher general | 1277 (29.9) | 781 (29.2) | 152 (31.5) | 298 (30.1) | 41 (38.3) | 5 (33.3) |
| Higher vocational, university | 975 (22.8) | 680 (25.4) | 89 (18.4) | 191 (19.3) | 11 (10.3) | 4 (26.7) |
| Smoking, n (%) |  |  |  |  |  |  |
| Never | 1348 (31.6) | 965 (36.1) | 130 (26.9) | 230 (23.3) | 21 (19.6) | 2 (13.3) |
| Former | 2152 (50.4) | 1276 (47.7) | 275 (56.9) | 541 (54.7) | 53 (49.5) | 7 (46.7) |
| Current | 749 (17.6) | 424 (15.9) | 77 (15.9) | 210 (21.2) | 33 (30.8) | 5 (33.3) |
| BMI, mean (SD) | 27.3 (4.1) | 27.3 (4.2) | 27.5 (3.8) | 27.3 (3.7) | 26.6 (3.5) | 27.8 (3.4) |
| Systolic blood pressure, mean (SD) | 142 (21.7) | 138 (20.7) | 147 (21.4) | 148 (22.1) | 150 (23.0) | 141 (18.2) |
| Diastolic blood pressure, mean (SD) | 82.9 (11.1) | 83.0 (10.7) | 83.1 (11.1) | 82.5 (11.9) | 81.4 (11.6) | 75.4 (8.3) |
| Total cholesterol, mean (SD) | 5.5 (1.1) | 5.6 (1.1) | 5.5 (1.1) | 5.5 (1.1) | 5.5 (1.2) | 5.0 (1.5) |
| HDL cholesterol, mean (SD) | 1.5 (0.4) | 1.5 (0.4) | 1.4 (0.4) | 1.4 (0.4) | 1.4 (0.4) | 1.2 (0.3) |
| eGFR, mean (SD) | 82.3 (14.8) | 84.0 (13.9) | 79.6 (15.0) | 79.2 (16.0) | 75.0 (17.7) | 65.8 (19.6) |
| Diabetes, n (%) | 588 (13.8) | 289 (10.8) | 82 (17.0) | 182 (18.4) | 31 (29.0) | 4 (26.7) |
| APOE-e4 carrier, n (%) | 1146 (26.9) | 706 (26.4) | 134 (27.7) | 279 (28.2) | 25 (23.4) | 2 (13.3) |
| Antithrombotic medication use, n (%) | 936 (21.9) | 391 (14.6) | 142 (29.4) | 334 (33.8) | 59 (55.1) | 10 (66.7) |
| Antihypertensive medication use, n (%) | 1683 (39.4) | 872 (32.6) | 236 (48.9) | 507 (51.3) | 59 (55.1) | 9 (60.0) |
| Lipid lowering medication use, n (%) | 1130 (26.5) | 572 (21.4) | 159 (32.9) | 343 (34.7) | 46 (43.0) | 10 (66.7) |
| History of (contralateral) stroke, n (%) | 44 (1.0) | 24 (0.9) | 9 (1.9) | 10 (1.0) | 1 (0.9) | 0 (0) |

Legend: SD = standard deviation; BMI = body mass index, in kg/m<sup>2</sup>; blood pressure in mmHg, cholesterol in mmol/l. Number of participants with missing data: education: 38 (0.9%), smoking: 18 (0.4%), BMI: 25 (0.6%), systolic blood pressure: 19 (0.4%), diastolic blood pressure: 19 (0.4%), total cholesterol: 83 (1.9%), HDL cholesterol: 84 (2.0%), eGFR: 348 (8.2%), APOE: 183 (4.3%), medication use: 13 (0.3%), diabetes: 8 (0.2%).

### Risk of dementia

During a total 41,251 person-years of follow-up (mean: 9.7 years), 311 (7.3%) participants developed dementia. Compared to individuals without carotid atherosclerosis, those with only plaques had an increased risk of dementia in age/sex-adjusted analyses (HR [95%CI]: 1.53 [1.17–2.01], Table S2), persistent after adjustment for cardiometabolic risk factors (HR: 1.39 [1.05–1.83], Figure 2). In contrast, we observed no significant effect of stenosis on dementia risk in age/sex-adjusted models (Table S2) nor in the fully adjusted models (Figure 2), with no clear trend by worsening degree of stenosis (1-49% stenosis, HR: 1.09 [0.85–1.40]; 50-99% stenosis, HR: 1.18 [0.70–1.99]; 70-99% stenosis, HR: 1.21 [0.44–3.35]). Only participants with a complete occlusion had a 4.6-fold increased risk of dementia (HR: 4.61 [1.83–11.66]; Figure 2). This high risk with occlusion drove up relative risk estimates of dichotomised stenosis, especially for ≥50% and ≥70% stenosis: when including occlusion, the HR for ≥50% stenosis was 1.27 (0.81–1.97) and for ≥70% 1.81 (0.91–3.59) but when excluding occlusion this dropped to 1.05 (0.64–1.73) for ≥50% stenosis and 1.07 (0.39–2.91) for ≥70% (Table S3).

**Figure 2:**
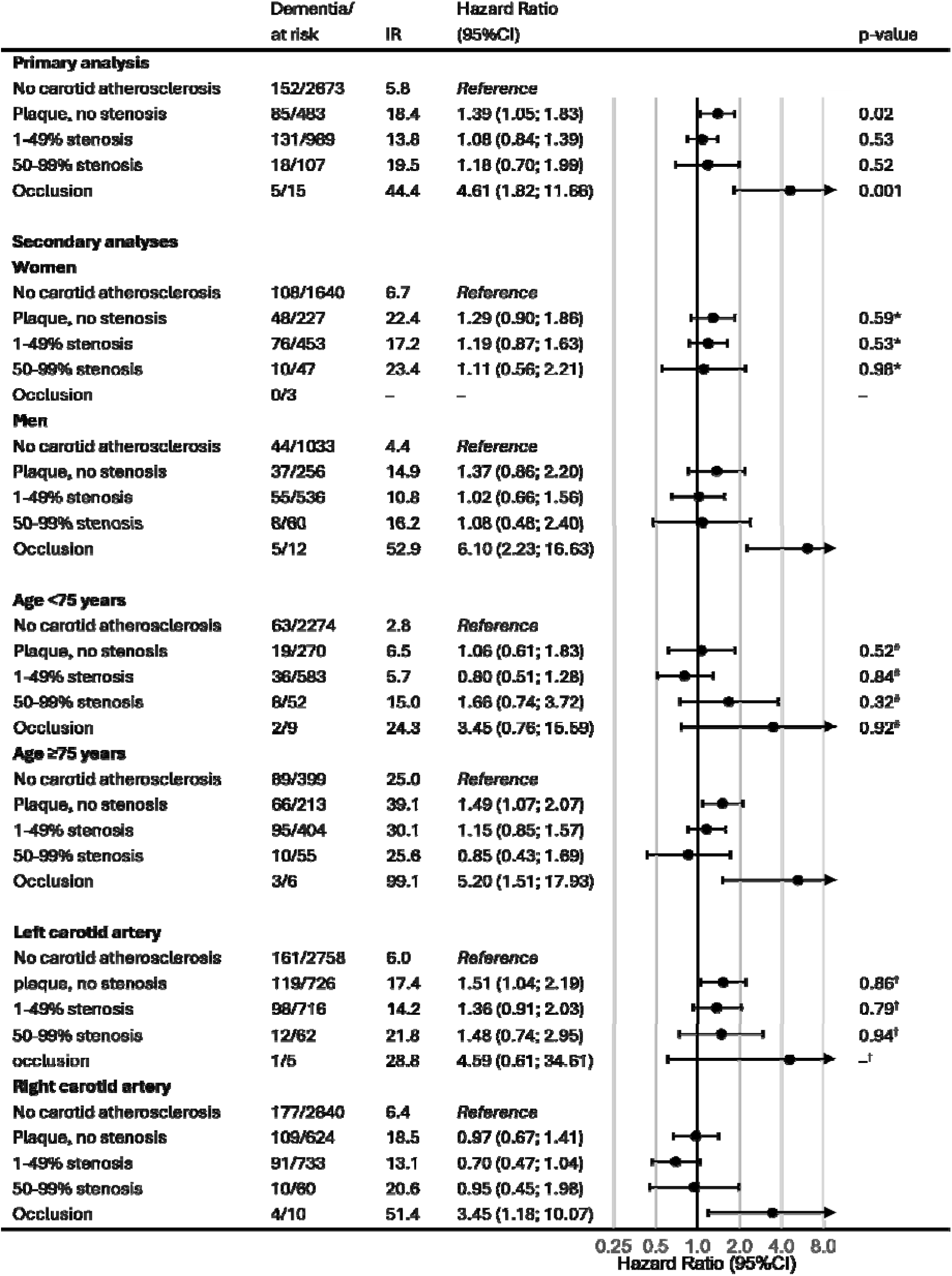
Effect of asymptomatic carotid artery atherosclerosis and stenosis on risk of dementia. Legend: Results from cox models adjusted for age, age, sex, smoking status, body mass index, systolic and diastolic blood pressure, total and HDL cholesterol, diabetes prevalence, eGFR, APOE-e4 carriership, use of antithrombotic, antihypertensive and lipid lowering medication, and a history of contralateral stroke. Left and right carotid artery variables were included in the same models. IR = incidence rate per 1000 person-years; HR = hazard ratio; CI = confidence interval, IMT = intima-media thickness (in mm). * = p-value for interaction with sex; # = p-value for interaction with age (continuous) in a model with also an interaction with age^2^, † = p-value for interaction with side (convergence error for occlusion).

When examining plaque characteristics on MRI, calcification tended to be associated with a moderate increase in dementia risk, whereas intraplaque haemorrhage and presence of a lipid core were not (Table 2). Among individuals with plaque, IMT did not increase dementia risk (HR per mm: 1.03 [0.87–1.22]).

**Table 2:** Effect of carotid plaque characteristics on risk of dementia, cognition and cognitive decline.

| **Dementia risk** | **Hazard Ratio (95%CI)** | **p-value** |
| --- | --- | --- |
| Intraplaque haemorrhage | 0.91 (0.67; 1.24) | 0.54 |
| Lipid necrotic core | 1.08 (0.82; 1.42) | 0.58 |
| Calcification | 1.40 (0.92; 2.12) | 0.11 |
| IMT (per mm) | 1.03 (0.87; 1.22) | 0.74 |
| **Cognition at baseline** | **β estimate (95%CI)** | **p-value** |
| Intercept | -0.239 (-0.775; 0.297) | 0.39 |
| Intraplaque haemorrhage | -0.076 (-0.160; 0.009) | 0.08 |
| Lipid necrotic core | 0.062 (-0.012; 0.136) | 0.10 |
| Calcification | -0.016 (-0.110; 0.079) | 0.75 |
| IMT (per mm) | -0.008 (-0.054; 0.038) | 0.75 |
| **Cognition over time** | **β estimate (95%CI)** | **p-value** |
| Time since baseline (years) | -0.078 (-0.095; -0.061) | <0.0001 |
| Intraplaque haemorrhage | 0.004 (-0.005; 0.014) | 0.38 |
| Lipid necrotic core | 0.005 (-0.003; 0.013) | 0.19 |
| Calcification | -0.009 (-0.017; 0.000) | 0.059 |
| IMT (per mm) | 0.000 (-0.005; 0.006) | 0.86 |

Legend: The cox model for dementia risk was adjusted for age, age<sup>2</sup>, sex, smoking status, body mass index, systolic and diastolic blood pressure, total and HDL cholesterol, diabetes prevalence, eGFR, APOE-e4 carriership, use of antithrombotic, antihypertensive and lipid lowering medication, and a history of contralateral stroke. The linear mixed model for cognition at baseline and cognition over time was adjusted for the same variables as the cox model and includes the interaction between age and time. Age was centred to improve interpretability of the intercept. CI = confidence interval, IMT = intimal medial thickness.

In secondary subgroup analyses (Figure 2), we observed no clear differences between men and women. Risk estimates for plaque without stenosis were somewhat higher in older participants than in younger participants (HR ≥75 years: 1.49 [1.07–2.07] vs. <75 years: 1.06 [0.61–1.83]), but an opposite age-trend was observed for ≥50% stenosis (HR ≥75 years: 1.66 [0.74–3.72]; and <75 years 0.85 [0.43–1.69]), and neither difference was statistically significant when formally testing for interaction. Risk estimates for dementia were numerically higher for left-sided than for right-sided atherosclerosis and stenosis, though interaction terms again were not statistically significant. Results were unchanged by censoring 196 participants who had ischemic stroke during follow-up did not change the risk of dementia in any of the stenosis categories.

### Cognitive assessment

Of all 4267 participants, 2961 (69.4%) participants completed at least two neuropsychological assessments, with a total of 8228 cognitive examinations during the study period (median: 2 [IQR: 1–3], Table S4). Participants with multiple examinations less often had carotid atherosclerosis, especially ≥50% stenosis, were younger, and had fewer cardiovascular risk factors (Table S5).

At baseline, participants with carotid atherosclerosis had lower scores on global cognition than those without atherosclerosis (Table S6), but in multivariable models this was attenuated and remained statistically significant only for occlusion (β: -0.503 [95% CI: -0.874; -0.131], Table 3). Global cognition declined over time by 0.074 SD annually (95% CI: 0.071-0.077), but this was unrelated to carotid atherosclerosis, with adjusted β for plaque without stenosis of -0.004 (-0.011; 0.004); for 1-49% stenosis -0.001 (-0.006; 0.005); for 50-99% stenosis: 0.008 (-0.009; 0.025); and for occlusion: 0.030 (-0.040; 0.099, Table 3). Age- and sex-adjusted analyses are presented in Table S6. When analysed as dichotomous exposure variables, individuals with carotid atherosclerosis, ≥50% stenosis, and ≥70% stenosis tended to have poorer global cognition at baseline, though these differences were not statistically significant and the nominal difference with ≥50% stenosis and ≥70% stenosis was driven by those with occlusion. Cognitive decline over time was not affected by carotid atherosclerosis, ≥50% and ≥70% stenosis in dichotomised analyses, either with or without occlusion (Table S3).

**Table 3:** Effect of asymptomatic carotid artery atherosclerosis and stenosis on cognition and cognitive decline.

| | $\beta$ estimate (95%CI) | p-value |
| --- | --- | --- |
| **Cognition at baseline** |  |  |
| Intercept | -0.049 (-0.346; 0.248) | 0.75 |
| No carotid atherosclerosis | *Reference* |  |
| Plaque, no stenosis | -0.047 (-0.116; 0.023) | 0.19 |
| 1-49% stenosis | -0.043 (-0.099; 0.012) | 0.13 |
| 50-99% stenosis | -0.122 (-0.264; 0.020) | 0.09 |
| Occlusion | -0.486 (-0.860; -0.112) | 0.01 |
| **Cognition over time** |  |  |
| Time since baseline (years) | -0.075 (-0.078; -0.072) | <0.0001 |
| No carotid atherosclerosis | *Reference* |  |
| Plaque, no stenosis | -0.003 (-0.010; 0.004) | 0.34 |
| 1-49% stenosis | -0.001 (-0.006; 0.005) | 0.84 |
| 50-99% stenosis | 0.008 (-0.009; 0.025) | 0.36 |
| Occlusion | 0.030 (-0.039; 0.099) | 0.40 |

Legend: The linear mixed model was adjusted for age, age<sup>2</sup>, sex, smoking status, body mass index, systolic and diastolic blood pressure, total and HDL cholesterol, diabetes prevalence, eGFR, APOE-e4 carriership, use of antithrombotic, antihypertensive and lipid lowering medication, and a history of contralateral stroke, and included an interaction term for age and time. Age was centred to improve interpretability of the intercept.

When examining plaque characteristics among individuals with carotid atherosclerosis, only calcification tended to be associated with a marginally faster decline in global cognition (Table 2). Neither calcification, intraplaque haemorrhage, nor lipid necrotic core were related to baseline global cognition (Table S7).

In secondary subgroup analyses, we observed no consistent differences in effect sizes by age, sex, or side of stenosis (all p_interaction_ >0.05, Table S8). Censoring participants with incident ischemic stroke also did not affect the results (Table S8).

Results for separate cognitive tests were broadly similar to those for global cognition (Table S7). Compared to people without carotid atherosclerosis, those with carotid atherosclerosis scored worse on tests at baseline, though this was only statistically significant for those with occlusion on the letter-digit substitution test (Table S8). Over time, individuals with plaque but no stenosis showed significantly steeper decline in the Stroop interference score than participants without carotid atherosclerosis (β per year: -0.015 [-0.028 to -0.002]). We did not find a significant effect of carotid atherosclerosis on any of the other cognitive test trajectories (Table S8).

### Mortality risk

Asymptomatic carotid artery stenosis was associated with small to moderate increases in mortality risk, with hazard ratios of 1.08 (95%CI: 0.89–1.45) for plaque without stenosis, 1.25 (1.08–1.45) for 1-49% stenosis, 1.35 (1.02–1.80) for 50-99% stenosis, and 1.73 (0.88–3.40) for complete occlusion (Table S9).

## DISCUSSION

In this population-based study with long-term follow-up of asymptomatic carotid artery stenosis, presence of plaque without stenosis and occlusion were associated with long-term dementia risk, but we did not observe a significant effect of stenosis on cognitive decline or dementia risk. The effect of carotid atherosclerosis on cognitive decline and dementia did not vary significantly with plaque characteristics on MRI.

In a concurrent systematic review and meta-analysis including results from this study, we found a significant association between ≥50% asymptomatic stenosis and dementia with a HR of 1.48 (95%CI: 1.06–2.07). However, that analysis included individuals with an occlusion. In this study, we showed that the relative risk estimates with ≥50% and ≥70% stenosis were strongly driven by those with an occlusion. This suggests that carotid stenosis only increases the risk of dementia when haemodynamics are severely impacted, as with occlusion. Previous studies indeed showed that cognition was most affected in people with stenosis and a reduced breath-holding index, which is a measure of cerebrovascular reserve, i.e. the capacity to increase cerebral blood flow in response to reduced perfusion pressure or metabolic demand. Furthermore, a meta-analysis showed that cerebrovascular reserve is reduced in only 29% of patients with severe ACAS,^28^ and we previously showed that in 21% of carotid arteries with ≥50% asymptomatic stenosis blood flow is not reduced at all.^10^ Together, this would explain why the 20-year follow-up post-hoc analysis of the ECST trial did not find an effect of carotid revascularisation on dementia.^29^

Whereas stenosis alone may not accelerate cognitive decline, our finding of an elevated dementia risk in individuals with plaque without stenosis underscores the role of carotid atherosclerosis in dementia aetiology. In this context, it is important to recognise that not all plaques induce luminal narrowing and that many undergo outward remodelling exclusively.^30^ Stenosis is therefore not an inevitable progression of the atherosclerotic process following plaque formation, which may explain why we did not observe an incrementally stronger association between atherosclerosis and dementia with increasing degrees of stenosis relative to its absence. Notably, we did find a nominally higher dementia risk and a trend toward faster cognitive decline in the presence of plaque calcification. Given that calcification occurs later in plaque development –reflecting a more prolonged disease process–these findings point to a prolonged atherosclerotic disease process as a contributor to dementia risk.^31^ ^32^ As such, cardiometabolic risk factor management remains the cornerstone to mitigate risk of both stroke and dementia.

Opposed to calcification, there was no signal that other plaque characteristics contribute to dementia risk, not even intra-plaque hemorrhage, which is a strong predictor of incident stroke.^18^ This suggests that the effect of carotid atherosclerosis is not mediated by ischemic stroke.^33^ Indeed, dementia diagnosis was preceded by incident ischemic stroke in only 1 out of 23 dementia cases of participants with ≥50% stenosis, and relative risks did not change materially when censoring participants at time of incident ischemic stroke. This also explains in part why risk increases in stroke with asymptomatic stenosis do not necessarily translate into cognitive decline and dementia. Unless future trials prove otherwise, additional cognitive benefit of carotid revascularisation in unselected ACAS patients is thus unlikely. The ongoing CREST-H trial aims to elucidate if revascularisation in ACAS patients with affected haemodynamics improves cognition.^34^ Though patients with carotid occlusion are not eligible for revascularisation, their observed increased risk of dementia may caution against overly strict blood pressure targets, in light of hemodynamic effects and previously described risk of orthostatic hypotension in this patient group.^35^

Strengths of this study include its large population-based sample with detailed phenotyping of stenosis degree and plaque characteristics on MRI, the meticulous long-term follow-up for cognitive decline and dementia, and the simultaneous analysis of carotid atherosclerosis with and without stenosis. As revascularisation of ACAS is rare in the Netherlands (0/107 participants with 50-99% stenosis in our sample), results provide a clear view on natural history under contemporary medical treatment. Certain limitations also need to be considered when interpreting the results. First, competing risk of mortality might lead to underestimation of the causal association of ACAS with dementia. However, this is unlikely to explain why we did not find a significant effect of ACAS on dementia, as we accounted for this in part in the Cox models and mortality risk was only moderately higher in people with stenosis than in those without. Second, attrition for dementia was low (2%) but substantially higher for detailed cognitive reassessment with the full test battery (29%), which may have led to underestimation of the association with cognitive decline. Third, despite the large overall sample, results were imprecise with respect to ≥70% stenosis and occlusion. Fourth, generalisability to non-White populations might be limited, for example in view of higher concurrent intracranial arterial disease in Asian populations.

In conclusion, asymptomatic carotid artery plaque and occlusion increase the risk of dementia, but we did not find a significant effect of ≥50% stenosis on cognitive decline or dementia. These results underscore the role of atherosclerosis in the aetiology of dementia, but do not support consideration of cognitive ability in clinical decision-making on treatment of ACAS.

## CONTRIBUTORS

C.V.J.B. was involved in conceptualisation, data curation, formal analysis, methodology, visualisation, writing – original draft, and writing – review and editing. A.C.P. was involved in formal analysis, writing – original draft, and writing – review and editing. Q.Y. was involved in writing – review and editing. M.K. was involved in conceptualisation, methodology, and writing – review and editing. A.L. was involved in conceptualisation, methodology, and writing – review and editing. M.K.I. was involved in conceptualisation, methodology, and writing – review and editing. D.B. was involved in conceptualisation, data curation, funding acquisition, methodology, supervision, and writing – review and editing. F.J.W. was involved in conceptualisation, data curation, funding acquisition, methodology, supervision, and writing – review and editing.

## DATA AVAILABILITY STATEMENT

Requests for the use of anonymised data can be directed to the management team of the Rotterdam Study, which has a protocol for approving data requests. Because of restrictions based on privacy regulations and informed consent of the participants, data cannot be made freely available in a public repository. D.B. and F.J.W. had full access to all study data and take responsibility for its integrity and the data analysis.

## ACKNOWLEDGEMENTS

The authors gratefully acknowledge the contribution of the study participants, general practitioners, and pharmacists of the Ommoord district who took part in the Rotterdam Study, as well as the contribution of the staff in facilitating data collection.

## REFERENCES

1. Howard DPJ, Gaziano L, Rothwell PM, Oxford Vascular S. Risk of stroke in relation to degree of asymptomatic carotid stenosis: a population-based cohort study, systematic review, and meta-analysis. Lancet Neurol 2021;20(3):193–202.

2. Bonati LH, Kakkos S, Berkefeld J, et al. European Stroke Organisation guideline on endarterectomy and stenting for carotid artery stenosis. Eur Stroke J 2021;6(2):I–XLVII.

3. Paraskevas KI, Mikhailidis DP, Ringleb PA, et al. An international, multispecialty, expert-based Delphi Consensus document on controversial issues in the management of patients with asymptomatic and symptomatic carotid stenosis. J Vasc Surg 2024;79(2):420–35 e1.

4. Force UPST, Krist AH, Davidson KW, et al. Screening for Asymptomatic Carotid Artery Stenosis: US Preventive Services Task Force Recommendation Statement. Jama 2021;325(5):476–81.

5. Donners SJA, van Velzen TJ, Cheng SF, et al. Optimised medical therapy alone versus optimised medical therapy plus revascularisation for asymptomatic or low-to-intermediate risk symptomatic carotid stenosis (ECST-2): 2-year interim results of a multicentre randomised trial. Lancet Neurol 2025;24(5):389–99.

6. Brott TG, Howard G, Lal BK, et al. Medical Management and Revascularization for Asymptomatic Carotid Stenosis. N Engl J Med 2026;394(3):219–31.

7. Brown MM, Bonati LH. Managing Asymptomatic Carotid Stenosis. N Engl J Med 2026;394(3):296–97.

8. Grotta JC. CREST-2: Providing Clarity on Managing Asymptomatic Carotid Stenosis. Stroke Vasc Interv Neurol 2026;6(2):e002266.

9. de Weerd M, Greving JP, Hedblad B, et al. Prevalence of asymptomatic carotid artery stenosis in the general population: an individual participant data meta-analysis. Stroke 2010;41(6):1294–7.

10. Wolters FJ, Vernooij MW, Roshchupkin GV, et al. Effect of carotid artery stenosis on cortical microinfarcts, white matter integrity, and brain volume: An interhemispheric comparison within the population-based Rotterdam Study. Cereb Circ Cogn Behav 2025;9:100391.

11. Debette S, Schilling S, Duperron M-G, et al. Clinical Significance of Magnetic Resonance Imaging Markers of Vascular Brain Injury: A Systematic Review and Meta-analysis. JAMA Neurology 2019;76(1):81–94. doi: 10.1001/jamaneurol.2018.3122

12. Wolters FJ, Zonneveld HI, Hofman A, et al. Cerebral Perfusion and the Risk of Dementia: A Population-Based Study. Circulation 2017;136(8):719–28.

13. Paraskevas KI, Brown MM, Lal BK, et al. Recent advances and controversial issues in the optimal management of asymptomatic carotid stenosis. J Vasc Surg 2024;79(3):695–703.

14. Crespo-Cuevas AM, Canento T, Hernández-Perez M, et al. The Barcelona-Asymptomatic Intracranial Atherosclerosis (AsIA) study: Subclinical cervico-cerebral stenosis and middle cerebral artery pulsatility index as predictors of long-term incident cognitive impairment. Atherosclerosis 2020;312:104–09.

15. Kitagawa K, Miwa K, Yagita Y, et al. Association between carotid stenosis or lacunar infarction and incident dementia in patients with vascular risk factors. Eur J Neurol 2015;22(1):187–92.

16. Naylor R, Rantner B, Ancetti S, et al. Editor’s Choice - European Society for Vascular Surgery (ESVS) 2023 Clinical Practice Guidelines on the Management of Atherosclerotic Carotid and Vertebral Artery Disease. Eur J Vasc Endovasc Surg 2023;65(1):7–111.

17. AbuRahma AF, Avgerinos ED, Chang RW, et al. Society for Vascular Surgery clinical practice guidelines for management of extracranial cerebrovascular disease. J Vasc Surg 2022;75(1S):4S–22S.

18. Bos D, Arshi B, van den Bouwhuijsen QJA, et al. Atherosclerotic Carotid Plaque Composition and Incident Stroke and Coronary Events. J Am Coll Cardiol 2021;77(11):1426–35.

19. Ikram MA, Kieboom BCT, Brouwer WP, et al. The Rotterdam Study. Design update and major findings between 2020 and 2024. Eur J Epidemiol 2024;39(2):183–206.

20. Iglesias del Sol A, Bots ML, Grobbee DE, et al. Carotid intima-media thickness at different sites: relation to incident myocardial infarction; The Rotterdam Study. Eur Heart J 2002;23(12):934–40.

21. van den Bouwhuijsen QJ, Vernooij MW, Hofman A, et al. Determinants of magnetic resonance imaging detected carotid plaque components: the Rotterdam Study. Eur Heart J 2012;33(2):221–9.

22. North American Symptomatic Carotid Endarterectomy Trial. Methods, patient characteristics, and progress. Stroke 1991;22(6):711–20.

23. Golden CJ. Identification of brain disorders by the Stroop Color and Word Test. J Clin Psychol 1976;32(3):654–8.

24. Welsh KA, Butters N, Mohs RC, et al. The Consortium to Establish a Registry for Alzheimer’s Disease (CERAD). Part V. A normative study of the neuropsychological battery. Neurology 1994;44(4):609–14.

25. Lezak MD. Neuropsychological assessment: Oxford University Press, USA 2004.

26. Lezak MD. Neuropsychological assessment in behavioral toxicology--developing techniques and interpretative issues. Scand J Work Environ Health 1984;10 Suppl 1:25–9.

27. Rietveld CA, Medland SE, Derringer J, et al. GWAS of 126,559 individuals identifies genetic variants associated with educational attainment. Science 2013;340(6139):1467–71.

28. Kamtchum-Tatuene J, Noubiap JJ, Wilman AH, et al. Prevalence of High-risk Plaques and Risk of Stroke in Patients With Asymptomatic Carotid Stenosis: A Meta-analysis. JAMA Neurol 2020;77(12):1524–35.

29. Halliday A, Sneade M, Björck M, et al. Editor’s Choice - Effect of Carotid Endarterectomy on 20 Year Incidence of Recorded Dementia: A Randomised Trial. Eur J Vasc Endovasc Surg 2022;63(4):535–45.

30. Schoenhagen P, Ziada KM, Vince DG, et al. Arterial remodeling and coronary artery disease: the concept of “dilated” versus “obstructive” coronary atherosclerosis. Journal of the American College of Cardiology 2001;38(2):297–306.

31. Stary HC, Chandler AB, Dinsmore RE, et al. A definition of advanced types of atherosclerotic lesions and a histological classification of atherosclerosis. A report from the Committee on Vascular Lesions of the Council on Arteriosclerosis, American Heart Association. Circulation 1995;92(5):1355–74.

32. Saba L, Nardi V, Cau R, et al. Carotid Artery Plaque Calcifications: Lessons From Histopathology to Diagnostic Imaging. Stroke 2022;53(1):290–97.

33. Paraskevas KI, Faggioli G, Ancetti S, Naylor AR. Editor’s Choice - Asymptomatic Carotid Stenosis and Cognitive Impairment: A Systematic Review. Eur J Vasc Endovasc Surg 2021;61(6):888–99.

34. Marshall RS, Lazar RM, Liebeskind DS, et al. Carotid revascularization and medical management for asymptomatic carotid stenosis - Hemodynamics (CREST-H): Study design and rationale. Int J Stroke 2018;13(9):985–91.

35. Starmans NLP, Wolters FJ, Leeuwis AE, et al. Orthostatic hypotension, cognition and structural brain imaging in hemodynamically impaired patients. J Neurol Sci 2024;461:123026.

