## Supplemental Material for "Asymptomatic carotid artery stenosis, cognitive decline and dementia: a prospective population-based cohort study"

### Supplements

#### Table S1: Strengthening the reporting of observational studies in epidemiology (STROBE) checklist

|  | **Item No** | **Recommendation** | **Page  No** |
| --- | --- | --- | --- |
| **Title and abstract** | 1 | (*a*) Indicate the study’s design with a commonly used term in the title or the abstract | 1/2 |
|  |  | (*b*) Provide in the abstract an informative and balanced summary of what was done and what was found | 2 |
| **Introduction** | | | |
| Background/rationale | 2 | Explain the scientific background and rationale for the investigation being reported | 4 |
| Objectives | 3 | State specific objectives, including any prespecified hypotheses | 4 |
| **Methods** | | | |
| Study design | 4 | Present key elements of study design early in the paper | 5 |
| Setting | 5 | Describe the setting, locations, and relevant dates, including periods of recruitment, exposure, follow-up, and data collection | 5-7 |
| Participants | 6 | (*a*) *Cohort study*—Give the eligibility criteria, and the sources and methods of selection of participants. Describe methods of follow-up | 5-7 |
|  |  | (*b*) *Cohort study*—For matched studies, give matching criteria and number of exposed and unexposed | n.a. |
| Variables | 7 | Clearly define all outcomes, exposures, predictors, potential confounders, and effect modifiers. Give diagnostic criteria, if applicable | 5-7 |
| Data sources/ measurement | 8 | For each variable of interest, give sources of data and details of methods of assessment (measurement). Describe comparability of assessment methods if there is more than one group | 5-7 |
| Bias | 9 | Describe any efforts to address potential sources of bias | 7-8 |
| Study size | 10 | Explain how the study size was arrived at | 5/21 |
| Quantitative variables | 11 | Explain how quantitative variables were handled in the analyses. If applicable, describe which groupings were chosen and why | 7-8 |
| Statistical methods | 12 | (*a*) Describe all statistical methods, including those used to control for confounding | 7-8 |
|  |  | (*b*) Describe any methods used to examine subgroups and interactions | 8 |
|  |  | (*c*) Explain how missing data were addressed | 7 |
|  |  | (*d*) *Cohort study*—If applicable, explain how loss to follow-up was addressed | 7 |
|  |  | (*e*) Describe any sensitivity analyses | 8 |
| **Results** | | | |
| Participants | 13* | (a) Report numbers of individuals at each stage of study—eg numbers potentially eligible, examined for eligibility, confirmed eligible, included in the study, completing follow-up, and analysed | 9, Fig1 |
|  |  | (b) Give reasons for non-participation at each stage | 9, Fig1 |
|  |  | (c) Consider use of a flow diagram | Fig1 |
| Descriptive data | 14* | (a) Give characteristics of study participants (eg demographic, clinical, social) and information on exposures and potential confounders | 9, Table1 |
|  |  | (b) Indicate number of participants with missing data for each variable of interest | Table1 |
|  |  | (c) *Cohort study*—Summarise follow-up time (eg, average and total amount) | 9 |
| Outcome data | 15* | *Cohort study*—Report numbers of outcome events or summary measures over time | 9 |
| Main results | 16 | (*a*) Give unadjusted estimates and, if applicable, confounder-adjusted estimates and their precision (eg, 95% confidence interval). Make clear which confounders were adjusted for and why they were included | Fig1, Tables 2, 3, S3, S5, S6, S9 |
|  |  | (*b*) Report category boundaries when continuous variables were categorized | 6, Table S7 |
|  |  | (*c*) If relevant, consider translating estimates of relative risk into absolute risk for a meaningful time period |  |
| Other analyses | 17 | Report other analyses done—eg analyses of subgroups and interactions, and sensitivity analyses | 9-10 |
| **Discussion** | | | |
| Key results | 18 | Summarise key results with reference to study objectives | 11 |
| Limitations | 19 | Discuss limitations of the study, taking into account sources of potential bias or imprecision. Discuss both direction and magnitude of any potential bias | 12-13 |
| Interpretation | 20 | Give a cautious overall interpretation of results considering objectives, limitations, multiplicity of analyses, results from similar studies, and other relevant evidence | 12 |
| Generalisability | 21 | Discuss the generalisability (external validity) of the study results | 13 |
| **Other information** | | | |
| Funding | 22 | Give the source of funding and the role of the funders for the present study and, if applicable, for the original study on which the present article is based | 13 |

#### Table S2: Crude and age/sex adjusted models for risk of dementia by carotid atherosclerosis categories

|  | **Crude model** | | **Age & sex adjusted model** | |
| --- | --- | --- | --- | --- |
|  | Hazard Ratio (95%CI) | p-value | Hazard Ratio (95%CI) | p-value |
| **Primary analysis** | | | | |
| No carotid atherosclerosis | *Reference* | | *Reference* | |
| Plaque, no stenosis | 3.04 (2.32; 3.97) | <0.0001 | 1.53 (1.17; 2.01) | 0.004 |
| <50% stenosis | 2.28 (1.80; 2.88) | <0.0001 | 1.21 (0.95; 1.54) | 0.12 |
| 50-99% stenosis | 3.36 (2.06; 5.47) | 0.001 | 1.45 (0.88; 2.38) | 0.14 |
| Occlusion | 7.91 (3.25; 19.30) | <0.0001 | 4.47 (1.82; 11.01) | 0.001 |
| **Additional analyses** |  |  |  |  |
| **Women** |  |  |  |  |
| No carotid atherosclerosis | *Reference* | | *Reference* | |
| Plaque, no stenosis | 3.22 (2.29; 4.54) | <0.0001 | 1.51 (1.07; 2.13) | 0.02 |
| <50% stenosis | 2.47 (1.84; 3.32) | <0.0001 | 1.28 (0.95; 1.73) | 0.10 |
| 50-99% stenosis | 3.43 (1.79; 6.57) | 0.0002 | 1.60 (0.83; 3.06) | 0.16 |
| Occlusion | – | – | – | – |
| **Men** |  |  |  |  |
| No carotid atherosclerosis | *Reference* | | *Reference* | |
| Plaque, no stenosis | 3.25 (2.09; 5.05) | <0.0001 | 1.55 (0.99; 2.43) | 0.055 |
| <50% stenosis | 2.36 (1.58; 3.53) | <0.0001 | 1.14 (0.76; 1.71) | 0.53 |
| 50-99% stenosis | 3.75 (1.76; 7.98) | 0.0006 | 1.25 (0.58; 2.69) | 0.57 |
| Occlusion | 12.46 (4.93; 31.48) | <0.0001 | 5.36 (2.11; 13.59) | 0.004 |
| **<75 year old** |  |  |  |  |
| No carotid atherosclerosis | *Reference* | | *Reference* | |
| Plaque, no stenosis | 2.03 (1.21; 3.40) | 0.008 | 1.30 (0.77; 2.20) | 0.33 |
| <50% stenosis | 1.79 (1.18; 2.71) | 0.006 | 1.14 (0.74; 1.74) | 0.56 |
| 50-99% stenosis | 4.86 (2.32; 10.17) | <0.0001 | 2.92 (1.39; 6.16) | 0.005 |
| Occlusion | 8.20 (2.00; 33.58) | <0.0001 | 3.61 (0.87; 14.99) | 0.08 |
| **≥75 year old** |  |  |  |  |
| No carotid atherosclerosis | *Reference* | | *Reference* | |
| Plaque, no stenosis | 1.58 (1.15; 2.17) | 0.002 | 1.56 (1.13; 2.15) | 0.007 |
| <50% stenosis | 1.23 (0.92; 1.64) | 0.15 | 1.22 (0.91; 1.63) | 0.19 |
| 50-99% stenosis | 1.11 (0.58; 2.14) | 0.75 | 1.02 (0.53; 1.96) | 0.96 |
| Occlusion | 5.05 (1.59; 16.00) | 0.006 | 5.33 (1.65; 17.18) | 0.005 |
| **Left carotid artery** | | | |  |
| No carotid atherosclerosis | *Reference* | | *Reference* | |
| plaque, no stenosis | 2.17 (1.45; 3.24) | 0.0002 | 1.56 (1.07; 2.27) | 0.02 |
| <50% stenosis | 1.82 (1.20; 2.75) | 0.005 | 1.49 (1.00; 2.22) | 0.051 |
| 50-99% stenosis | 2.50 (1.24; 5.02) | 0.01 | 1.73 (0.88; 3.41) | 0.11 |
| Occlusion | 4.08 (0.55; 30.15) | 0.17 | 3.55 (0.48; 26.20) | 0.21 |
| **Right carotid artery** |  |  |  |  |
| No carotid atherosclerosis | *Reference* | | *Reference* | |
| Plaque, no stenosis | 1.52 (1.02; 2.28) | 0.04 | 1.00 (0.69; 1.46) | 0.98 |
| <50% stenosis | 1.11 (0.74; 1.68) | 0.61 | 0.74 (0.50; 1.10) | 0.14 |
| 50-99% stenosis | 1.84 (0.89; 3.80) | 0.10 | 0.98 (0.48; 2.00) | 0.94 |
| Occlusion | 4.03 (1.38; 11.78) | 0.01 | 3.15 (1.09; 9.10) | 0.03 |

Legend: Results from Cox models. The age and sex adjusted model includes age^2^. CI = confidence interval; IMT = intima-media thickness (in mm). Left and right carotid artery variables were included in the same models.

#### Table S3: Effect of carotid atherosclerosis, ≥50% and ≥70 stenosis on risk of dementia, cognition and cognitive decline, assessed as dichotomous exposures including and excluding occlusion

|  | **Including occlusion** | | **Excluding occlusion** | |
| --- | --- | --- | --- | --- |
| **Dementia risk** | Hazard Ratio (95%CI) | p-value | Hazard Ratio (95%CI) | p-value |
| Carotid atherosclerosis vs no carotid atherosclerosis | 1.20 (0.96; 1.50) | 0.10 | 1.19 (0.96; 1.49) | 0.12 |
| ≥50% stenosis vs <50% stenosis | 1.27 (0.81; 1.97) | 0.29 | 1.05 (0.64; 1.73) | 0.83 |
| ≥70% stenosis vs <70% stenosis | 1.81 (0.91; 3.59) | 0.08 | 1.07 (0.39; 2.91) | 0.90 |
| **Cognition at baseline** | β estimate (95%CI) | p-value | β estimate (95%CI) | p-value |
| Carotid atherosclerosis vs no carotid atherosclerosis | -0.046 (-0.094; 0.002) | 0.06 | -0.042 (-0.090; 0.005) | 0.08 |
| ≥50% stenosis vs <50% stenosis | -0.118 (-0.249; 0.012) | 0.07 | -0.071 (-0.209; 0.068) | 0.32 |
| ≥70% stenosis vs <70% stenosis | -0.197 (-0.424; 0.030) | 0.09 | -0.032 (-0.317; 0.252) | 0.83 |
| **Cognition over time** | β estimate (95%CI) | p-value | β estimate (95%CI) | p-value |
| Carotid atherosclerosis vs no carotid atherosclerosis | -0.001 (-0.006; 0.003) | 0.64 | -0.001 (-0.006; 0.003) | 0.61 |
| ≥50% stenosis vs <50% stenosis | 0.010 (-0.007; 0.026) | 0.25 | 0.008 (-0.009; 0.025) | 0.33 |
| ≥70% stenosis vs <70% stenosis | 0.025 (-0.016; 0.065) | 0.23 | 0.024 (-0.026; 0.074) | 0.35 |

Legend: Cox models for dementia risk are adjusted for age, age^2^, sex, smoking status, body mass index, systolic and diastolic blood pressure, total and HDL cholesterol, diabetes prevalence, eGFR, APOE-e4 carriership, use of antithrombotic, antihypertensive and lipid lowering medication, and a history of contralateral stroke. Linear mixed models for cognition at baseline and cognition over time are adjusted for the same variables as the cox model and includes the interaction between age and time. CI = confidence interval.

#### Table S4: Number of cognitive testing rounds, stratified by exposure category

|  | **Number of participants (%)** | | | | |
| --- | --- | --- | --- | --- | --- |
| Number of cognitive testing rounds | Without carotid atherosclerosis (N = 2673) | Plaque, no stenosis  (N = 483) | 1-49% stenosis  (N = 989) | 50-99% stenosis (N = 107) | Occlusion (N = 15) |
| 0 | 4 (0.1) | 15 (3.1) | 61 (6.2) | 8 (7.5) | 1 (6.7) |
| 1 | 742 (27.8) | 140 (29.9) | 279 (30.1) | 47 (47.5) | 9 (64.3) |
| 2 | 1348 (50.5) | 165 (35.3) | 330 (35.6) | 28 (28.3) | 4 (28.6) |
| 3 | 576 (21.6) | 163 (34.8) | 319 (34.4) | 24 (24.2) | 1 (7.1) |
| 4 | 3 (0.1) | 0 (0) | 0 (0) | 0 (0) | 0 (0) |

Legend: N = number of participants.

#### Table S5. Baseline characteristics of those with only one cognitive examination vs those with multiple

|  | **Single examination**  **(N = 1217)** | **Multiple examinations  (N = 2961)** |
| --- | --- | --- |
| Age, mean (SD) | 69.7 (11.2) | 66.3 (8.90) |
| Female sex, n (%) | 679 (55.8) | 1645 (55.6) |
| Education, n (%) |  |  |
| Primary only | 157 (12.9) | 209 (7.1) |
| Lower/intermediate general, lower vocational | 479 (39.4) | 1095 (37.7) |
| Intermediate vocational, higher general | 357 (29.3) | 900 (30.4) |
| Higher vocational, university | 224 (18.4) | 757 (25.6) |
| Smoking, n (%) |  |  |
| Never | 359 (29.5) | 970 (32.8) |
| Former | 595 (48.9) | 1520 (51.3) |
| Current | 263 (21.6) | 471 (15.9) |
| BMI, mean (SD) | 27.2 (4.1) | 27.3 (4.0) |
| Systolic blood pressure, mean (SD) | 143 (22.7) | 141 (21.1) |
| Diastolic blood pressure, mean (SD) | 82.7 (11.3) | 83.1 (10.9) |
| Total cholesterol, mean (SD) | 5.4 (1.1) | 5.6 (1.1) |
| HDL cholesterol, mean (SD) | 1.5 (0.4) | 1.5 (0.4) |
| eGFR, mean (SD) | 79.8 (15.5) | 83.2 (13.8) |
| Diabetes, n (%) | 185 (15.2) | 383 (12.9) |
| APOE-e4 carrier, n (%) | 335 (27.5) | 813 (27.5) |
| Antithrombotic medication use, n (%) | 334 (27.4) | 562 (19.0) |
| Antihypertensive medication use, n (%) | 557 (45.8) | 1082 (36.5) |
| Lipid lowering medication use, n (%) | 351 (28.8) | 754 (25.5) |
| History of (contralateral) stroke, n (%) | 27 (2.2) | 16 (0.5) |
| Carotid atherosclerosis, n (%) |  |  |
| Without carotid atherosclerosis | 742 (61.0) | 1927 (65.1) |
| Plaque, no stenosis | 140 (11.5) | 328 (11.1) |
| 1-49% stenosis | 279 (22.9) | 649 (21.9) |
| 50-99% stenosis | 47 (3.9) | 52 (1.8) |
| Occlusion | 9 (0.7) | 5 (0.2) |

Legend: SD = standard deviation; BMI = body mass index, in kg/m2; blood pressure in mmHg, cholesterol in mmol/l. Missingness was imputed using the average of 5-fold multiple imputation with the “mice” R package, carotid atherosclerosis was complete since it was the population that was selected for the main analyses.

#### Table S6: Crude and age/sex adjusted model for effect of asymptomatic carotid artery atherosclerosis and stenosis on cognition and cognitive decline

|  | **Crude model** | | **Age & sex adjusted model** | |
| --- | --- | --- | --- | --- |
|  | β estimate (95%CI) | p-value | β estimate (95%CI) | p-value |
| **Baseline** |  |  |  |  |
| Intercept | 0.444 (0.411; 0.477) | <0.0001 | 0.270 (0.227; 0.314) | <0.0001 |
| No carotid atherosclerosis | *Reference* | | *Reference* | |
| Plaque, no stenosis | -0.517 (-0.602; -0.432) | <0.0001 | -0.079 (-0.153; -0.005) | 0.04 |
| <50% stenosis | -0.501 (-0.566; -0.436) | <0.0001 | -0.079 (-0.153; -0.005) | 0.0004 |
| 50-99% stenosis | -0.801 (-0.976; -0.626) | <0.0001 | -0.220 (-0.369; -0.071) | 0.004 |
| Occlusion | -1.093 (-1.560; -0.627) | <0.0001 | -0.559 (-0.954; -0.165) | 0.006 |
| **Over time** |  |  |  |  |
| Time since baseline (years) | -0.066 (-0.069; -0.064) | <0.0001 | -0.074 (-0.077; -0.071) | <0.0001 |
| No carotid atherosclerosis | *Reference* | | *Reference* | |
| Plaque, no stenosis | -0.020 (-0.027; -0.012) | <0.001 | -0.003 (-0.010; 0.004) | 0.35 |
| <50% stenosis | -0.013 (-0.019; -0.008) | <0.0001 | -0.001 (-0.006; 0.005) | 0.83 |
| 50-99% stenosis | -0.008 (-0.026; 0.010) | 0.39 | -0.001 (-0.006; 0.005) | 0.39 |
| Occlusion | -0.009 (-0.083; 0.066) | 0.82 | 0.019 (-0.050; 0.089) | 0.59 |

Legend: Age/sex-adjusted model includes age^2^ and the interaction between age and time. Age was centred to improve interpretability of the intercept.

#### Table S7: Crude and age/sex adjusted models for the effects of carotid plaque characteristics on risk of dementia, cognition and cognitive decline

|  | **Crude model** | | **Age & sex adjusted model** | |
| --- | --- | --- | --- | --- |
| **Dementia risk** | Hazard Ratio (95%CI) | p-value | Hazard Ratio (95%CI) | p-value |
| Intraplaque haemorrhage | 1.18 (0.88; 1.59) | 0.27 | 0.94 (0.69; 1.28) | 0.70 |
| Lipid necrotic core | 1.03 (0.79; 1.34) | 0.85 | 1.12 (0.86; 1.47) | 0.40 |
| Calcification | 2.06 (1.38; 3.05) | 0.0004 | 1.45 (0.97; 2.18) | 0.07 |
| IMT (per mm) | 0.98 (0.84; 1.15) | 0.82 | 0.99 (0.83; 1.17) | 0.88 |
| **Cognition at baseline** | β estimate (95%CI) | p-value | β estimate (95%CI) | p-value |
| Intercept | 0.208 (0.007; 0.409) | 0.04 | 0.241 (0.061; 0.421) | 0.01 |
| Intraplaque haemorrhage | -0.274 (-0.379; -0.169) | <0.0001 | -0.072 (-0.163; 0.018) | 0.12 |
| Lipid necrotic core | 0.073 (-0.019; 0.165) | 0.12 | 0.111 (0.033; 0.189) | 0.005 |
| Calcification | -0.285 (-0.401; -0.169) | <0.0001 | -0.052 (-0.151; 0.048) | 0.31 |
| IMT (per mm) | -0.016 (-0.073; 0.042) | 0.60 | -0.018 (-0.067; 0.031) | 0.57 |
| **Cognition over time** | β estimate (95%CI) | p-value | β estimate (95%CI) | p-value |
| Time since baseline (years) | -0.077 (-0.095; -0.059) | <0.0001 | -0.077 (-0.094; -0.060) | <0.0001 |
| Intraplaque haemorrhage | -0.003 (-0.014; 0.007) | 0.51 | 0.004 (-0.005; 0.014) | 0.37 |
| Lipid necrotic core | 0.002 (-0.007; 0.010) | 0.70 | 0.005 (-0.003; 0.013) | 0.20 |
| Calcification | -0.020 (-0.030; -0.011) | <0.0001 | -0.009 (-0.018; -0.000) | 0.048 |
| IMT (per mm) | 0.002 (-0.004; 0.007) | 0.55 | 0.000 (-0.005; 0.005) | 0.92 |

Legend: Results from the age/sex-adjusted cox model for dementia risk including age^2^. Results from the linear mixed model for cognition at baseline and cognition over time including age^2^ and the interaction between age and time. Age was centred to improve interpretability of the intercept. CI = confidence interval, IMT = intimal medial thickness.

#### Table S8: Effect of asymptomatic carotid artery atherosclerosis and stenosis on cognition and cognitive decline, subgroup analyses

|  | **β-estimate (95% confidence interval)** | | | | | | |
| --- | --- | --- | --- | --- | --- | --- | --- |
|  | Men | Women | <75 years | ≥75 years | Left carotid artery | Right carotid artery | Censoring incident stroke |
| **Cognition at baseline** |  |  |  |  |  |  |  |
| Intercept | -0.129 (-0.576; 0.319) | 0.513 (0.104; 0.922) | 0.182 (-0.151; 0.514) | 0.185 (-0.627; 0.997) | 0.107 (-0.191; 0.404) | | 0.111 (-0.185; 0.407) |
| No carotid atherosclerosis | *Reference* | *Reference* | *Reference* | *Reference* | *Reference* | *Reference* | *Reference* |
| Plaque, no stenosis | -0.081 (-0.176; 0.014) | -0.005 (-0.107; 0.096) | -0.016 (-0.101; 0.069) | -0.083 (-0.210; 0.043) | -0.072 (-0.166; 0.021) | 0.059 (-0.038; 0.155) | -0.042 (-0.111; 0.027) |
| 1-49% stenosis | -0.043 (-0.120; 0.033) | -0.045 (-0.124; 0.034) | -0.017 (-0.080; 0.047) | -0.079 (-0.189; 0.031) | -0.078 (-0.174; 0.018) | 0.031 (-0.066; 0.127) | -0.036 (-0.091; 0.018) |
| 50-99% stenosis | -0.072 (-0.255; 0.110) | -0.116 (-0.336; 0.105) | -0.106 (-0.296; 0.083) | -0.100 (-0.325; 0.125) | -0.103 (-0.306; 0.099) | 0.019 (-0.180; 0.218) | -0.099 (-0.239; 0.042) |
| Occlusion | -0.503 (-0.896; -0.110) | 0.107 (-2.402; 2.615) | -0.646 (-1.090; -0.201) | -0.321 (-1.034; 0.392) | -0.387 (-0.985; 0.212) | -0.534 (-1.043; -0.026) | -0.505 (-0.875; -0.135) |
| **Cognition over time** |  |  |  |  |  |  |  |
| Time since baseline (years) | -0.072 (-0.077; -0.068) | -0.075 (-0.080; -0.071) | -0.074 (-0.077; -0.071) | -0.152 (-0.251; -0.054) | -0.074 (-0.077; -0.071) | | -0.074 (-0.077; -0.070) |
| No carotid atherosclerosis | *Reference* | *Reference* | *Reference* | *Reference* | *Reference* | *Reference* | *Reference* |
| Plaque, no stenosis | 0.003 (-0.006; 0.012) | -0.012 (-0.022; -0.001) | -0.002 (-0.010; 0.005) | -0.005 (-0.026; 0.016) | 0.005 (-0.005; 0.014) | -0.012 (-0.021; -0.002) | -0.003 (-0.010; 0.004) |
| 1-49% stenosis | -0.003 (-0.010; 0.004) | 0.002 (-0.006; 0.010) | -0.001 (-0.007; 0.004) | 0.006 (-0.012; 0.023) | 0.012 (0.003; 0.022) | -0.012 (-0.022; -0.002) | -0.001 (-0.006; 0.005) |
| 50-99% stenosis | 0.017 (-0.005; 0.038) | -0.004 (-0.030; 0.023) | 0.011 (-0.008; 0.030) | -0.004 (-0.048; 0.041) | 0.016 (-0.007; 0.040) | -0.004 (-0.029; 0.021) | 0.009 (-0.008; 0.027) |
| Occlusion | 0.026 (-0.039; 0.092) | 3.344 (-7.874; 14.562) | 0.029 (-0.042; 0.100) | 0.078 (-0.160; 0.315) | 0.034 (-0.052; 0.120) | 0.037 (-0.084; 0.157) | 0.029 (-0.040; 0.098) |

Legend: Results from linear mixed models, adjusted for age, age^2^, sex (except in the stratified by sex models), smoking status, body mass index, systolic and diastolic blood pressure, total and HDL cholesterol, diabetes prevalence, eGFR, APOE-e4 carriership, use of antithrombotic, antihypertensive and lipid lowering medication, and a history of contralateral stroke, and included an interaction term for age and time. Right and left sided stenosis was included in the same model. Age was centred to improve interpretability of the intercept. P-values for interaction terms carotid atherosclerosis category with sex and carotid atherosclerosis category with age were all > 0.05.

|  | **LDST** | | **Stroop 3** | | **WFT** | | **WLTdel** | | **PPB** | |
| --- | --- | --- | --- | --- | --- | --- | --- | --- | --- | --- |
|  | β estimate  (95% CI) | p-value | Β estimate  (95% CI) | p-value | Β estimate  (95% CI) | p-value | Β estimate  (95% CI) | p-value | β estimate  (95% CI) | p-value |
| **Baseline** |  |  |  |  |  |  |  |  |  |  |
| Intercept | 0.027 (-0.326; 0.381) | 0.88 | -0.078 (-0.413; 0.257) | 0.65 | -0.208 (-0.581; 0.165) | 0.27 | 0.087 (-0.261; 0.435) | 0.62 | 0.544 (0.257; 0.832) | 0.0002 |
| No carotid atherosclerosis | *Reference* | | *Reference* | | *Reference* | | *Reference* | | *Reference* | |
| Plaque, no stenosis | -0.068 (-0.152; 0.016) | 0.11 | -0.037 (-0.115; 0.040) | 0.35 | 0.023 (-0.069; 0.114) | 0.63 | -0.029 (-0.114; 0.057) | 0.51 | -0.029 (-0.100; 0.042) | 0.42 |
| 1-49% stenosis | -0.049 (-0.116; 0.017) | 0.15 | 0.007 (-0.054; 0.068) | 0.82 | -0.019 (-0.091; 0.053) | 0.61 | -0.071 (-0.139; -0.004) | 0.04 | -0.009 (-0.065; 0.047) | 0.74 |
| 50-99% stenosis | -0.117 (-0.289; 0.054) | 0.18 | -0.058 (-0.217; 0.101) | 0.48 | -0.059 (-0.246; 0.128) | 0.54 | -0.056 (-0.231; 0.118) | 0.53 | -0.045 (-0.191; 0.101) | 0.54 |
| Occlusion | -0.615 (-1.065; -0.164) | 0.008 | -0.332 (-0.759; 0.094) | 0.13 | -0.335 (-0.833; 0.162) | 0.19 | -0.299 (-0.767; 0.170) | 0.21 | -0.289 (-0.681; 0.104) | 0.15 |
| **Over time** |  |  |  |  |  |  |  |  |  |  |
| Time since baseline (years) | -0.049 (-0.052; -0.045) | <0.0001 | -0.034 (-0.040; -0.028) | <0.0001 | -0.030 (-0.034; -0.025) | <0.0001 | -0.058 (-0.063; -0.053) | <0.0001 | -0.107 (-0.111; -0.103) | <0.0001 |
| No carotid atherosclerosis | *Reference* | | *Reference* | | *Reference* | | *Reference* | | *Reference* | |
| Plaque, no stenosis | 0.001 (-0.006; 0.009) | 0.71 | -0.015 (-0.028; -0.002) | 0.02 | -0.008 (-0.018; 0.003) | 0.14 | 0.001 (-0.010; 0.011) | 0.88 | 0.006 (-0.003; 0.014) | 0.20 |
| 1-49% stenosis | -0.003 (-0.009; 0.003) | 0.29 | -0.005 (-0.014; 0.005) | 0.33 | -0.004 (-0.012; 0.004) | 0.31 | 0.004 (-0.004; 0.012) | 0.34 | 0.006 (-0.000; 0.013) | 0.06 |
| 50-99% stenosis | 0.001 (-0.018; 0.019) | 0.93 | 0.013 (-0.018; 0.044) | 0.41 | -0.007 (-0.032; 0.017) | 0.57 | 0.016 (-0.009; 0.040) | 0.22 | 0.015 (-0.006; 0.035) | 0.16 |
| Occlusion | 0.014 (-0.062; 0.091) | 0.71 | -0.005 (-0.118; 0.108) | 0.93 | 0.019 (-0.079; 0.118) | 0.70 | 0.027 (-0.073; 0.127) | 0.60 | 0.083 (-0.001; 0.167) | 0.052 |

#### Table S9: Effect of asymptomatic carotid artery atherosclerosis and stenosis on different cognitive test results

Legend: Results are from linear mixed models, adjusted for age, age^2^, sex, smoking status, body mass index, systolic and diastolic blood pressure, total and HDL cholesterol, diabetes prevalence, eGFR, APOE-e4 carriership, use of antithrombotic, antihypertensive and lipid lowering medication, a history of contralateral stroke, and the interaction between age and time. Age was centred to improve interpretability of the intercept. All scores were standardised and Stroop 3 scores inverted to ensure higher scores represent better outcome. LDST = letter digit substitution test; WFT = word fluency test; WLTdel = Word learning test – delayed recall; PPB = Purdue pegboard Test.

#### Table S10: Effect of asymptomatic carotid artery atherosclerosis and stenosis on risk of mortality

|  |  |  | **Crude model** | | **Age & sex adjusted model** | | **Fully adjusted model** | |
| --- | --- | --- | --- | --- | --- | --- | --- | --- |
|  | Died/ at risk | MR | HR (95%CI) | p-value | HR (95%CI) | p-value | HR (95%CI) | p-value |
| No carotid atherosclerosis | 398/ 2673 | 14.8 | *Reference* | | *Reference* | | *Reference* | |
| Plaque, no stenosis | 175/ 483 | 35.4 | 2.24 (1.88; 2.68) | <0.0001 | 1.14 (0.95; 1.36) | 0.17 | 1.08 (0.89; 1.30) | 0.44 |
| 1-49% stenosis | 406/ 989 | 41.0 | 2.61 (2.27; 3.00) | <0.0001 | 1.39 (1.21; 1.61) | <0.0001 | 1.25 (1.08; 1.45) | 0.003 |
| 50-99% stenosis | 61/ 107 | 62.3 | 4.09 (3.12; 5.35) | <0.0001 | 1.76 (1.34; 2.32) | 0.0001 | 1.35 (1.02; 1.80) | 0.04 |
| Occlusion | 9/ 15 | 67.4 | 4.61 (2.38; 8.93) | <0.0001 | 2.29 (1.18; 4.46) | 0.01 | 1.73 (0.88; 3.40) | 0.11 |

Legend: Results from cox models, age & sex adjusted includes age^2^, fully adjusted model is adjusted for age, age^2^, sex, smoking status, body mass index, systolic and diastolic blood pressure, total and HDL cholesterol, diabetes prevalence, eGFR, APOE-e4 carriership, use of antithrombotic, antihypertensive and lipid lowering medication, and a history of contralateral stroke. MR = mortality rate per 1000 person-years, HR = hazard ratio, CI = confidence interval.

#### Figure S1: Proportional hazards assumption

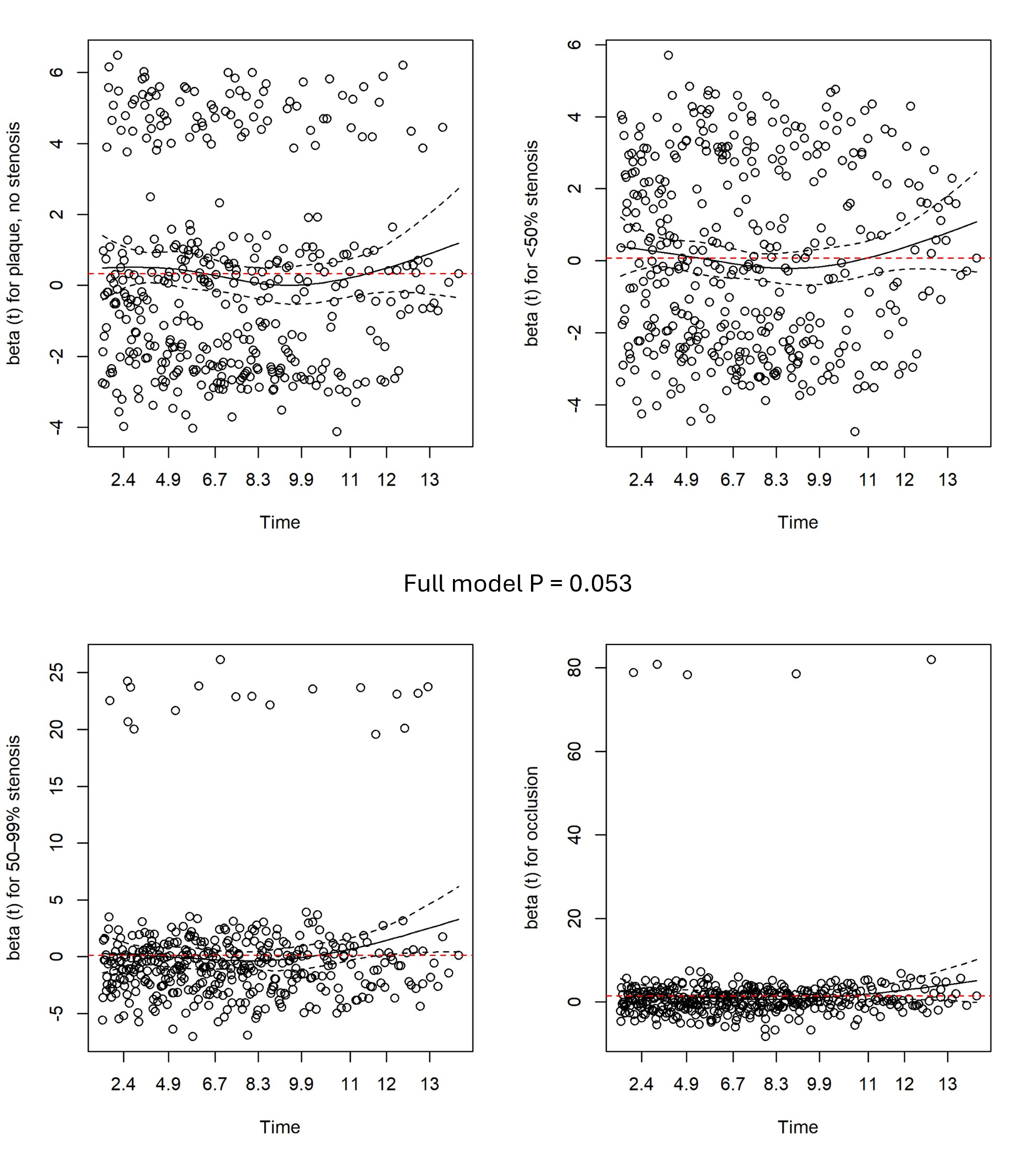

Legend: Graphs show the scaled Schoenfeld residuals over time of the fully adjusted model for the effect of carotid atherosclerosis on dementia risk. P values are for tests of the proportional hazards assumptions of the full model and individual coefficients. Time is in years. Dashed red lines indicate the coefficient’s betas.

#### Figure S2: Linear mixed model assumptions

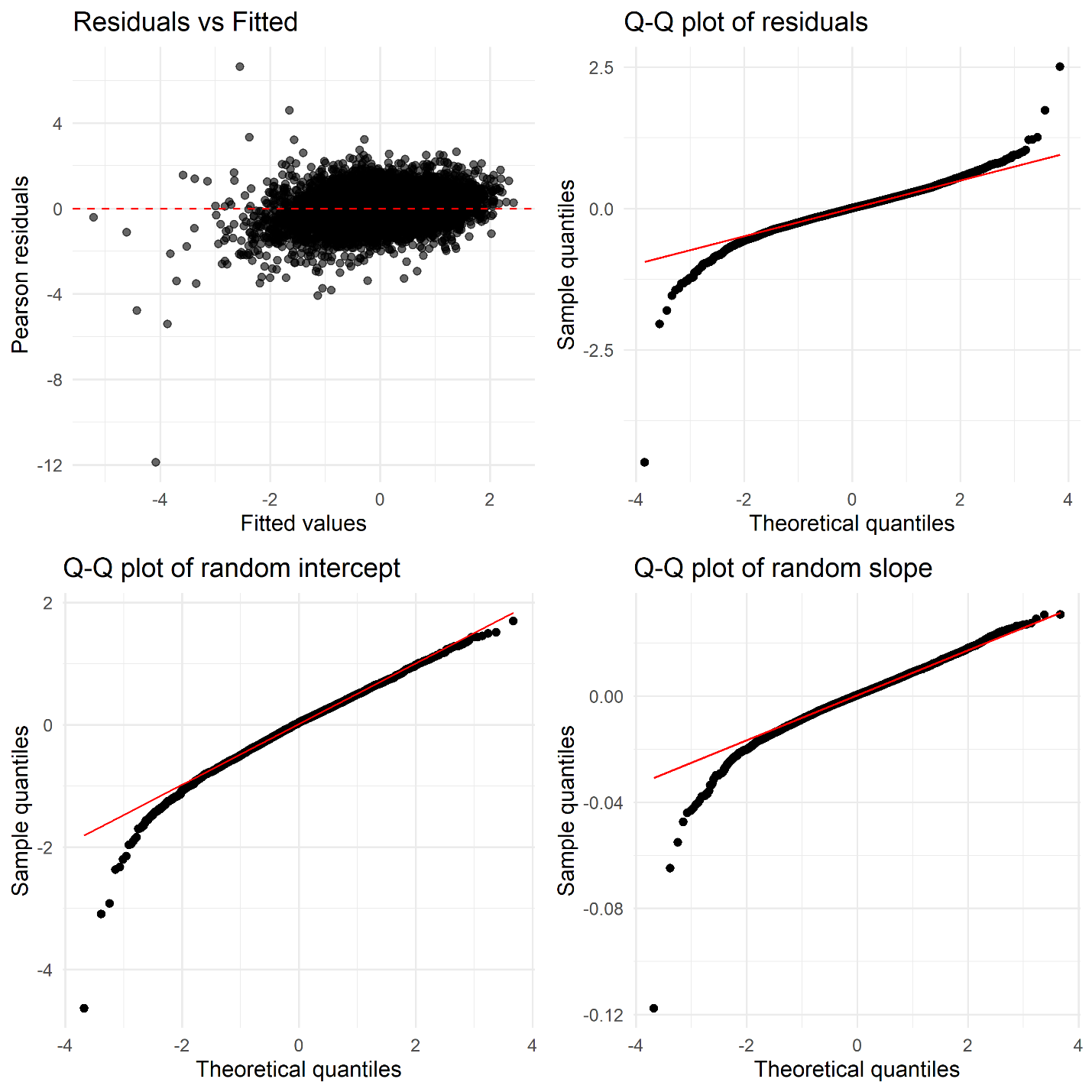

Legend: Plots are QQ-plots and standardised residuals plot of the fully adjusted linear mixed model for global cognition.

#### **Figure S3: Age-specific prevalence of asymptomatic carotid artery stenosis**

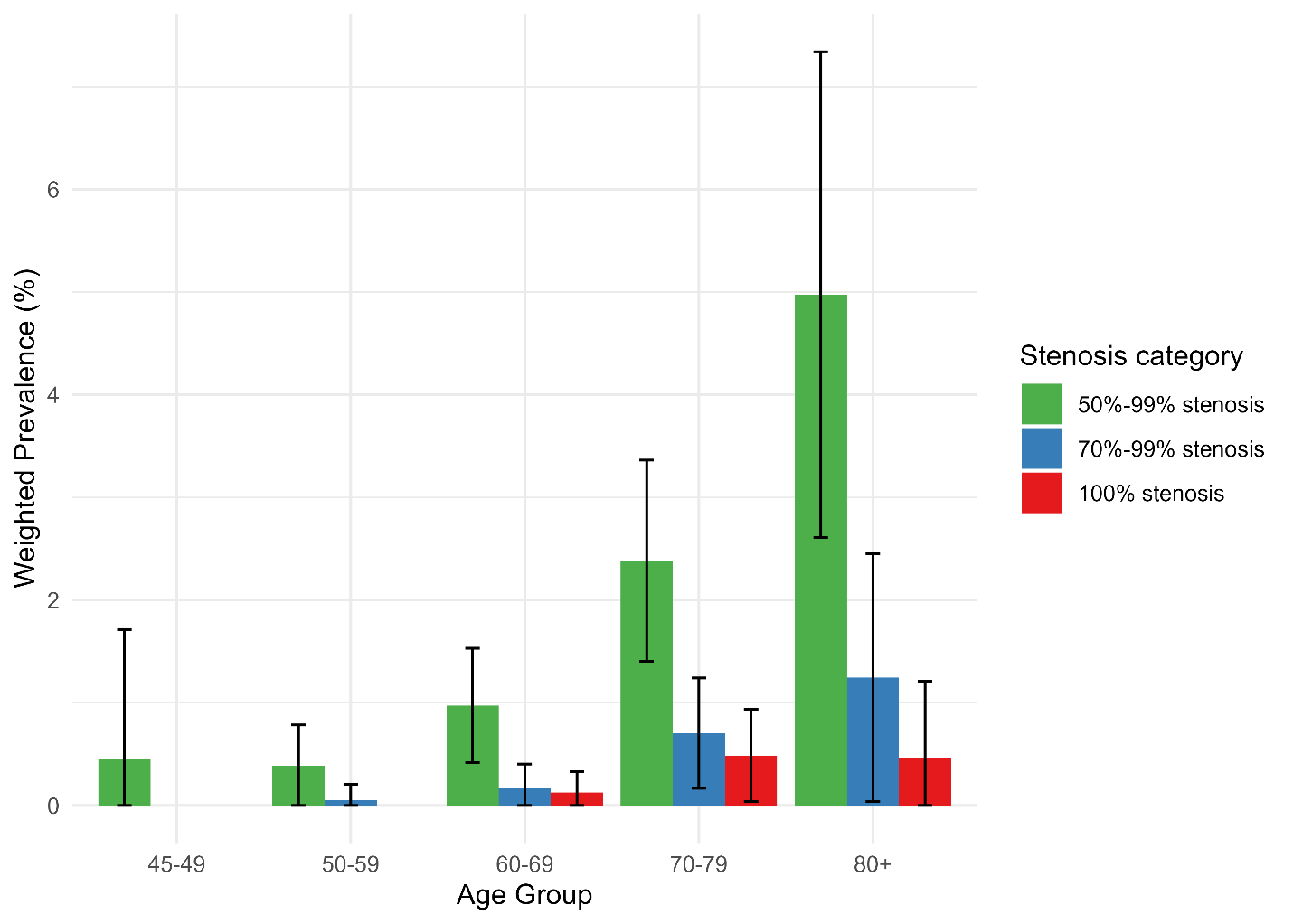

| Age, y | | 45-49 | | 50-59 | 60-69 | 70-79 | 80+ |
| --- | --- | --- | --- | --- | --- | --- | --- |
| n total | | 116 | | 995 | 1398 | 1272 | 486 |
| n per stenosis severity | | | | |  |  |  |
| 50-99% | 1 | | 7 | | 23 | 44 | 32 |
| 70-99% | 0 | | 1 | | 4 | 13 | 8 |
| 100% | 0 | | 0 | | 3 | 9 | 3 |
| Weighted prevalence in % (95%CI) | | | | | | | |
| 50-99% | | 0.5 (0.0; 1.7) | | 0.4 (0; 0.8) | 1.0 (0.4; 1.5) | 2.4 (1.4; 3.4) | 5.0 (2.6; 7.4) |
| 70-99% | | 0 | | 0.1 (0; 0.2) | 0.2 (0.0; 0.4) | 0.7 (0.2; 1.2) | 1.3 (0.04; 2.5) |
| 100% | | 0 | | 0 | 0.1 (0.0; 0.3) | 0.5 (0.05; 0.9) | 0.5 (0.0; 1.2) |

Legend: Prevalences are weighted by the probability of being invited for the carotid MRI for those with IMT >2.5mm, which was 50.6% of the total sample with IMT >2.5mm. y = years, n = number of participants. Error bars represent 95% confidence interval. The 95% confidence interval for prevalence was calculated using the normal approximation method.
